# Capsid integrity quantitative PCR to determine virus infectivity in environmental and food applications – a systematic review

**DOI:** 10.1101/2020.05.08.20095364

**Authors:** Mats Leifels, Cheng Dan, Emanuele Sozzi, David C. Shoults, Stefan Wuertz, Skorn Mongkolsuk, Kwanrawee Sirikanchana

**Affiliations:** Singapore Centre for Life Sciences Engineering, Nanyang Technological University (NTU), Singapore; Gilling’s School of Global Public Health, Department of Environmental Science and Engineering, University of North Carolina at Chapel Hill, NC, USA; Civil and Resource Engineering, Dalhousie University, Halifax, Nova Scotia, Canada; School of Civil and Environmental Engineering, NTU, Singapore; Research Laboratory of Biotechnology, Chulabhorn Research Institute, Bangkok, Thailand; Center of Excellence on Environmental Health and Toxicology, CHE, Ministry of Education, Bangkok, Thailand

**Keywords:** (6) azo dye, EMA, PMA, microbial contamination, viability, water quality

## Abstract

Capsid-integrity quantitative PCR (qPCR), a molecular detection method for infectious viruses combining azo-dye pretreatment with qPCR, has been widely used in recent years; however, variations in pretreatment conditions for various virus types can limit the efficacy of specific protocols. By identifying and critically synthesizing forty-two recent peer-reviewed studies employing capsid-integrity qPCR for viruses in the last decade (2009 to 2019) in the fields of food safety and environmental virology, we aimed to establish recommendations for the detection of infectious viruses. Intercalating dyes are effective measures of viability in PCR assays provided the viral capsid is damaged; viruses that have been inactivated by other causes, such as loss of attachment or genomic damage, are less well detected using this approach. Although optimizing specific protocols for each virus is recommended, we identify a framework for general assay conditions. These include concentrations of ethidium monoazide, propidium monoazide or its derivates between 10 and 200 µM; incubation on ice or at room temperature (20 - 25°C) for 5 to 120 min; and dye activation using LED or high light (500 – 800 Watts) exposure for periods ranging from 5 to 20 min. These simple steps can benefit the investigation of infectious virus transmission in routine (water) monitoring settings and during viral outbreaks such as the current COVID-19 pandemic or endemic diseases like dengue fever.

**Graphical abstract:** 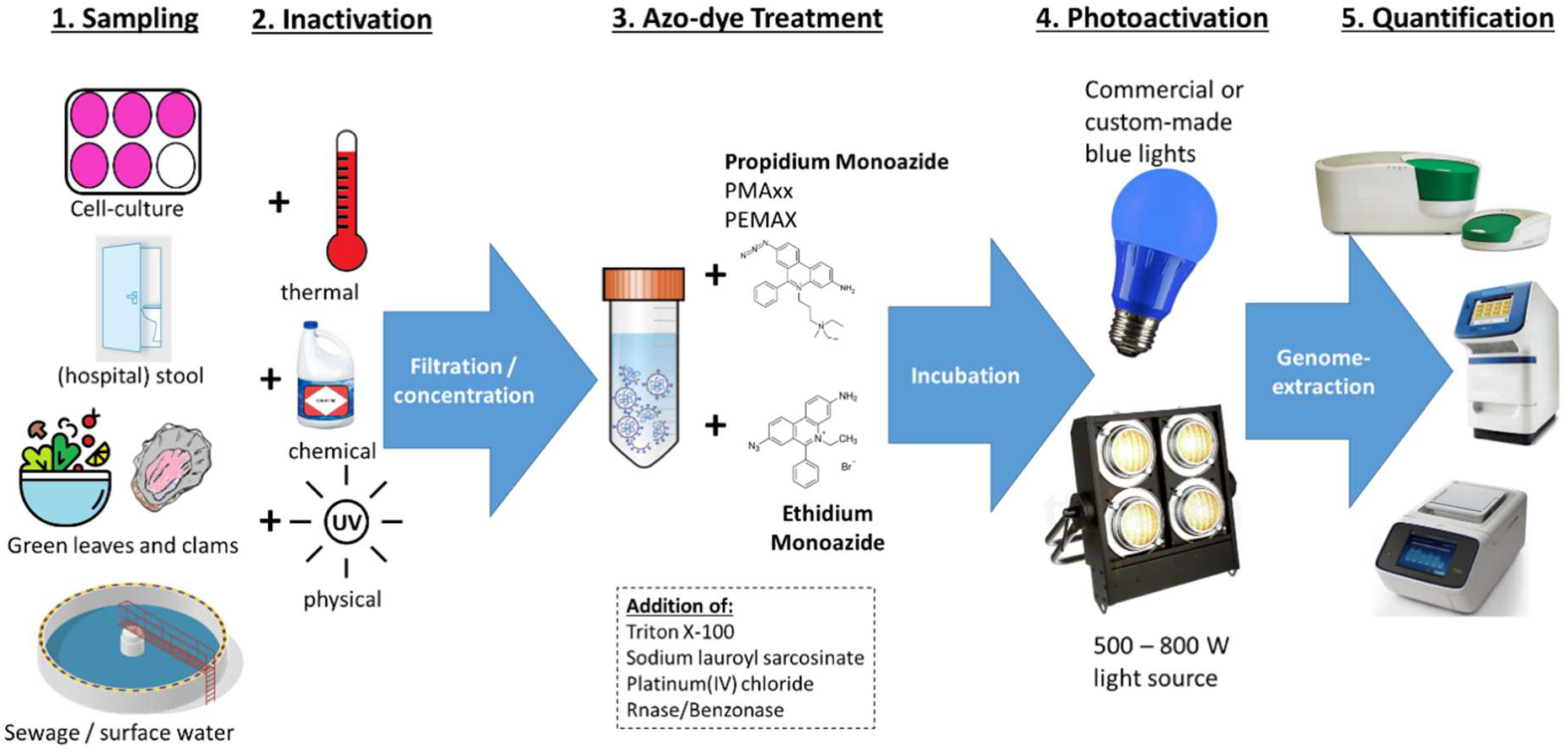

## 1. Introduction

Global population expansion and climate change are poised to increase both freshwater demand and wastewater production. Exposure to waterborne and foodborne pathogens through recreational activities, irrigation water, and food consumption, as well as associated occupations, poses a risk to public health in high and low resource environments (Efstratiou et al., 2017; Gibson, 2014). To date, more than 150 enteric viruses have been described to cause waterborne-associated human illnesses, including gastrointestinal and chronic infections (Sinclair et al., 2009). In addition, enteric viruses have shown a significantly higher persistence in the aquatic environment compared to conventional fecal indicator bacteria (Rames et al., 2016). Several enteric viruses relevant to human health could pass conventional sewage treatment in high numbers, thus posing a health risk when partially treated reclaimed sewage is utilized to irrigate fruits and vegetables (Brouwer et al., 2018) or released into the aquatic environment of rivers and lakes (Hellmér et al., 2014). Consequently, viral infectivity measurements are included in guidelines of water reuse for potable and non-potable purposes to demonstrate water reuse safety and evaluate water treatment efficiencies through log-reduction value achievements (Farkas et al., 2020; Gerba and Betancourt, 2019). Due to their high infectivity and transmission rate as well as usually relatively low infectious dose, virus analysis in water and on fomites is frequently used when investigating likelihood of water-/surface borne transmission. This is especially the case in viral outbreaks causing acute and chronic illnesses like the Ebola virus disease, severe acute respiratory syndrome (SARS), Middle Eastern respiratory syndrome (Mers), seasonal dengue outbreaks in (sub-)tropical regions and the current COVID-19 pandemic caused by SARS-Coronavirus-2 (Corman et al., 2020; Grubaugh et al., 2019)).

While still the gold standard, culture-based methods for propagation of infectious human pathogenic viruses in a laboratory environment require specialized facilities and experienced personnel and test results may only become available after five to ten days (Rodriguez et al., 2009). Molecular techniques using quantitative polymerase chain reaction (qPCR) are faster and have been successfully used in the past two decades to determine virus loads in the aquatic environment and to comply with food safety regulations (Bosch et al., 2018; Gerba and Betancourt, 2019). While robust, cost-efficient and uniquely sensitive and specific, qPCR has the severe limitation of not being able to differentiate between infectious and non-infectious virus particles, thus overestimating the number of viruses present in a sample (Chhipi-Shrestha et al., 2017). Novel approaches like modifying the targeted gene sequence or the length of the PCR product, or amplifying less stable messenger RNA after reverse transcription to DNA (e.g. (Ho et al., 2016; Ko et al., 2003; Polston et al., 2014; Wu et al., 2019)) generally lack robustness and sensitivity.

One of the most established qPCR modifications to measure infectivity is capsid integrity qPCR, an approach where samples are pre-treated with the intercalating azo dyes propidium monoazide (PMA), ethidium monoazide (EMA) or their derivates PMAxx and PEMAX. First described almost two decades ago by Nogva et al. (2003) to allow the identification of viable but non-cultivable bacteria, this technique has been successfully adapted to remove putatively false-positive qPCR signals deriving from virions with broken capsids in complex matrices like sewage and surface water (Leifels et al., 2016; Randazzo et al., 2018a). Based on the principle that an azo dye can only enter virions with a damaged capsid to covalently and irreversibly bind with viral DNA or RNA, this pretreatment can block amplification of nucleic acids due to the detachment of the polymerase when it encounters the dye-genome complex. Subsequently, only genomic targets are amplified that originate from intact virions while those nucleic acids that are free (outside the virion) or belong to non-infectious viruses are removed from the quantitative PCR. This indirect viable measurement method has been especially useful for those viruses for which cell cultivation-based detection has been difficult, but has yet to be fully validated (Estes et al., 2019). One known limitation of the azo intercalating dyes is their inability to differentiate viruses that have lost their infectivity due to damaged nucleic acids but whose capsid remains intact, a condition often found after UV-C treatment (Leifels et al., 2015). Moreover, there are numerous factors that affect the efficacy of a method, including virus type, inactivation method, type of dye and its concentration. Incubation conditions and light source are also crucial in the applicability of capsid integrity qPCR as reflected by the great range of capsid integrity pretreatment conditions in the literature. Consequently, the objectives of this review were to evaluate the efficiency of azo dye pretreatment conditions as stated in current literature and to establish protocols and considerations of the capsid integrity qPCR methods for virus infectivity monitoring.

## 2. Literature Search and Analysis Strategy

The guidelines for systematic article search and selection as recommended in the PRISMA Statement have been adopted in this work (Moher et al., 2009; Moher et al., 2015). To ensure reproducibility, a search string was constructed in accordance with the Cochrane Handbook (Cochrane, 2019), and a search was conducted in March 2020 (Figure 1) in relevant databases like *Pubmed, Scopus, Ovid, Medline* and *Web of Knowledge* to cover relevant articles in English since the first introduction of the azo dye pretreatment in 2003 (Supplementary Table S1). Articles were screened according to specific criteria.

**Figure 1:**
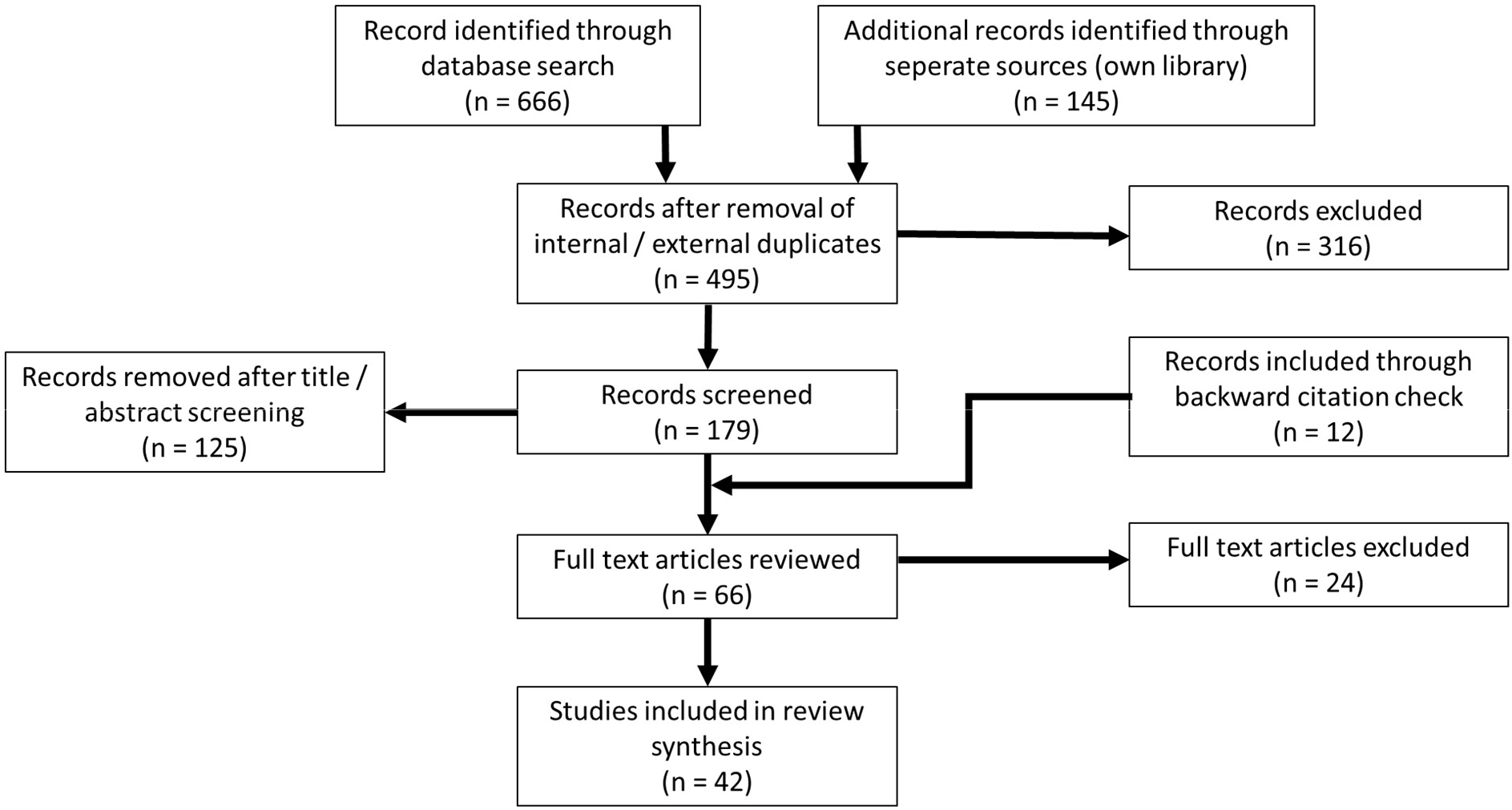
PRISMA flowchart of the literature search strategy and the number of included and excluded articles

For quality control and to follow the recommendations of Cochrane (2019), two of the authors (M.L. and K.S.) conducted the title and abstract evaluation in parallel, with a third author (E.S.) acting as tie breaker in the case of disagreement. As depicted in Figure 2, the 42 articles represented here include studies discussing the application of azo dyes (PMA, EMA, PMAxx and PEMAX) to determine virus or bacteriophage infectivity in the context of food safety or environmental virology as well as comparisons of azo dye applications with other established methods such as cell-culture, phage plaque assay or conventional qPCR (Supplementary Table S2).

**Figure 2:**
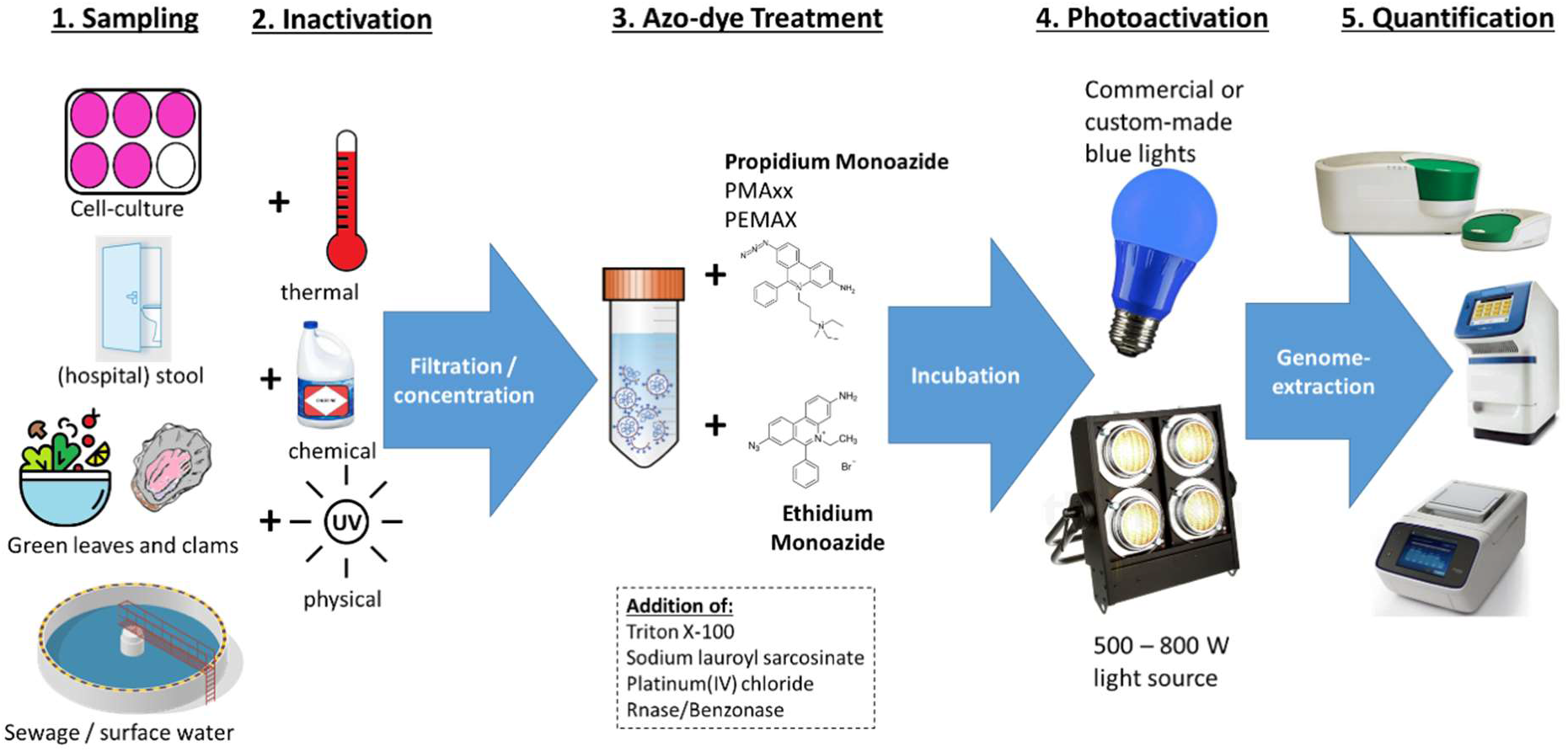
Schematic overview of a typical case study structure. One or more virus samples (taken from the environment or a culture collection) are split in two and one portion is inactivated, the other not, before filtration and concentration steps. Azo-dye pretreatment is then conducted under various incubation conditions and concentrations of PMA, PMAxx, PEMAX or EMA, either in the presence or absence of additives like surfactants and enzymes, before the tubes are exposed to light for photo activation. While early studies used high-energy light sources (500 – 800 Watt) to initiate the formation of the light induced dye-genome complex, more recent articles have focused on low energy LED in the blue light spectrum. Virus quantification is done with qPCR or ddPCR for quantitative, or endpoint PCR for qualitative, detection after genome extraction.

Disinfection methods utilized in water treatment or food safety have also used capsid integrity qPCR to determine the efficiency in virus inactivation (Lee et al., 2018; Leifels et al., 2015; Randazzo et al., 2018b).

## 3. Systematic Review Results

### 3.1 Viruses studied

All known types of enteric viruses, whether they contain single- or double-stranded DNA or RNA, with or without an envelope, have been studied with capsid-integrity qPCR methods (Figure 3).

**Figure 3:**
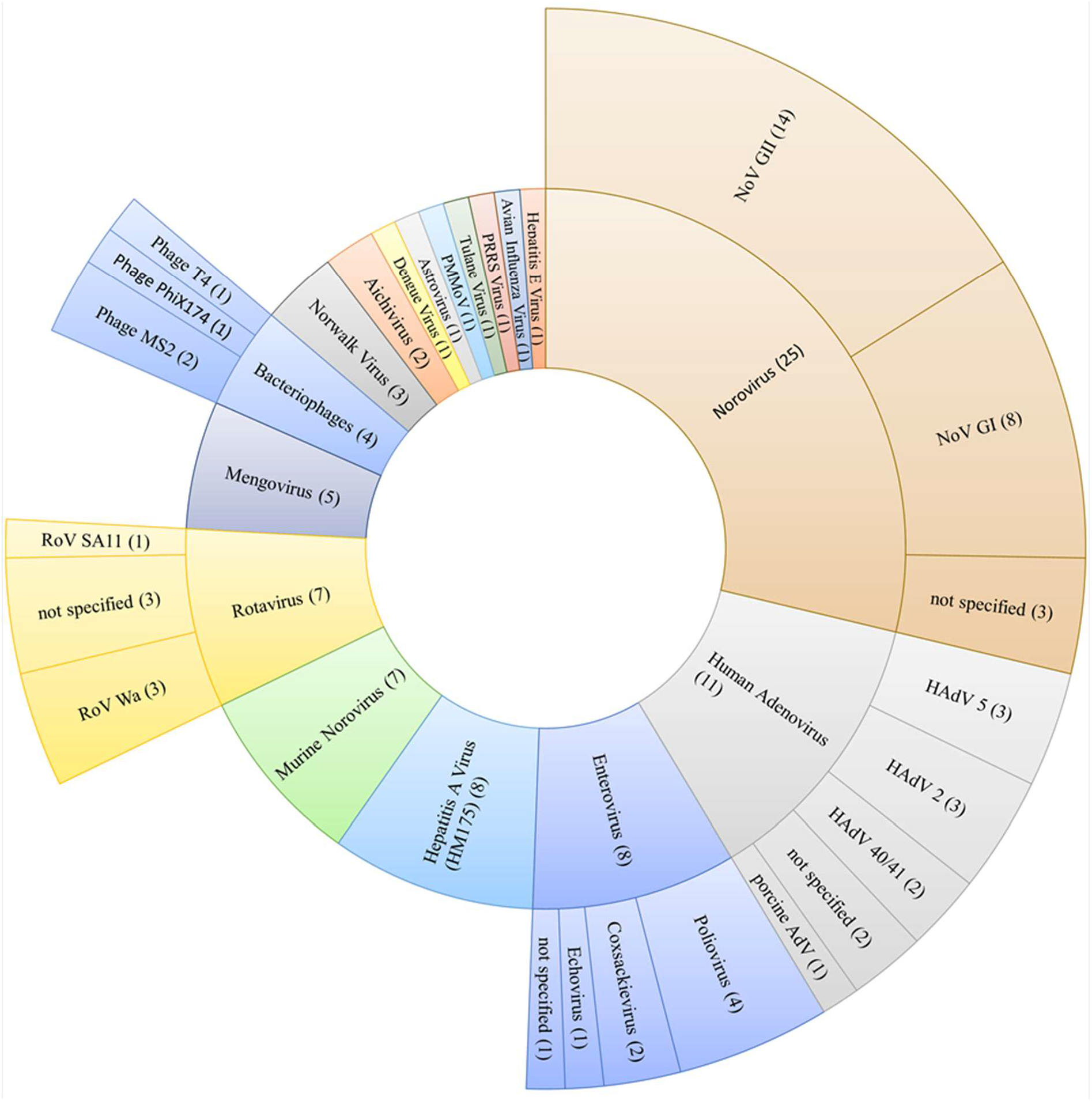
Genus and strain number of viruses investigated and frequency of occurrence in case studies (in parentheses). In cases where no strain name was listed, the virus is referred to as ‘not specified’.

The majority of studies investigated two or more viruses (69%) with an emphasis on the detection of norovirus GI and GII (NoV GI/NoV GII) together with hepatitis A virus (HAV). The increase in the number of studies on capsid integrity of NoV’s and HAV between 2016 and 2020 is most likely associated with the release of ISO/TS 15216-2:2013 (ISO, 2013) that regulates the quantification of NoV and HAV using qPCR, and subsequently has been replaced with an updated version (ISO, 2019). The risk associated with norovirus outbreaks due to a low infectious dose and high rate of particle shedding by infected individuals, together with the absence of commercially available animal tissue cell lines, also explains the interest in developing azo dye pretreatments for the detection of NoV (Blanco et al., 2017; Lowther et al., 2019). Murine norovirus (MNV) and mengovirus, two viruses without relevance to human health, have been chosen as both are widely used as process control surrogates because they lack human pathogenicity and are easy to propagate on certain cell lines. Of the articles included, only three cover bacteriophages infecting coliform bacteria as their host (phages MS2, PhiX174 and T4). As the laboratory safety requirements, as well as workload for culture-based detection, of those phages are significantly lower than for enteric viruses, the necessity to establish alternative methods to determine phage infectivity is not as urgent as for enteric viruses (Toribio-Avedillo et al., 2020). Their increasing relevance in the context of microbial source tracking will likely lead to more studies in the future (Ogilvie et al., 2018).

The intercalating dye infectivity assay was successfully applied to most viruses tested, with few exceptions. Bacteriophage T4 infecting *Escherichia coli* required higher temperatures for inactivation than other viruses, with extremely high heat (110°C) for significant capsid damage; lower temperatures (85°C) and proteolysis were not effective (Fittipaldi et al., 2012). Moreover, capsid disruption of MNV was more challenging than for other viruses in the same studies, i.e., HAdV, poliovirus, rotavirus, and bacteriophages phiX174 and MS2, using heat treatment (Kim and Ko, 2012; Leifels et al., 2015). Lack of efficacy of an azo dye method with avian influenza virus was suspected to be due to natural characteristics of an enveloped virus that make it difficult for EMA to penetrate the compromised capsid (Graiver et al., 2010). However, the PMA dye assay was successfully applied to detect dengue virus, another enveloped, single-stranded RNA virus (Huang et al., 2016), lending further support to the conclusion that PMA is more effective than EMA in removing false positive signals in qPCR (Fittipaldi et al., 2012; Gedalanga and Olson, 2009; Leifels et al., 2019).

## 4. Origin of studied viruses

The viruses analyzed were split evenly between strains obtained from culture-collections and wild types, those obtained from clinical, environmental and food samples (Table 1). While progress has been made in introducing protocols for their propagation in the laboratory, an established cell line to propagate NoV GI/GII has yet to arrive (Estes et al., 2019; Veronica et al., 2018). Subsequently, the studies included in this review analyzed NoV in stool samples obtained from hospitals, in sewage or in surface water. One study evaluated the presence of NoV GI/GII in struvite, a phosphate source used for fertilization, that was reclaimed from wastewater sludge (Yee et al., 2019). A limitation of all studies on clinical, environmental and food samples is the absence of information regarding absolute quantification of infectious and non-infectious viruses to evaluate the assay performance. Other complicating factors include process recovery loss and inhibition effects in qPCR assays that appeared to affect RNA viruses more than DNA viruses regardless of azo-dye pretreatment (Leifels et al., 2016). Next we discuss the assay conditions that influence the efficacy of capsid integrity qPCR in measuring infective viruses.

**Table 1:**
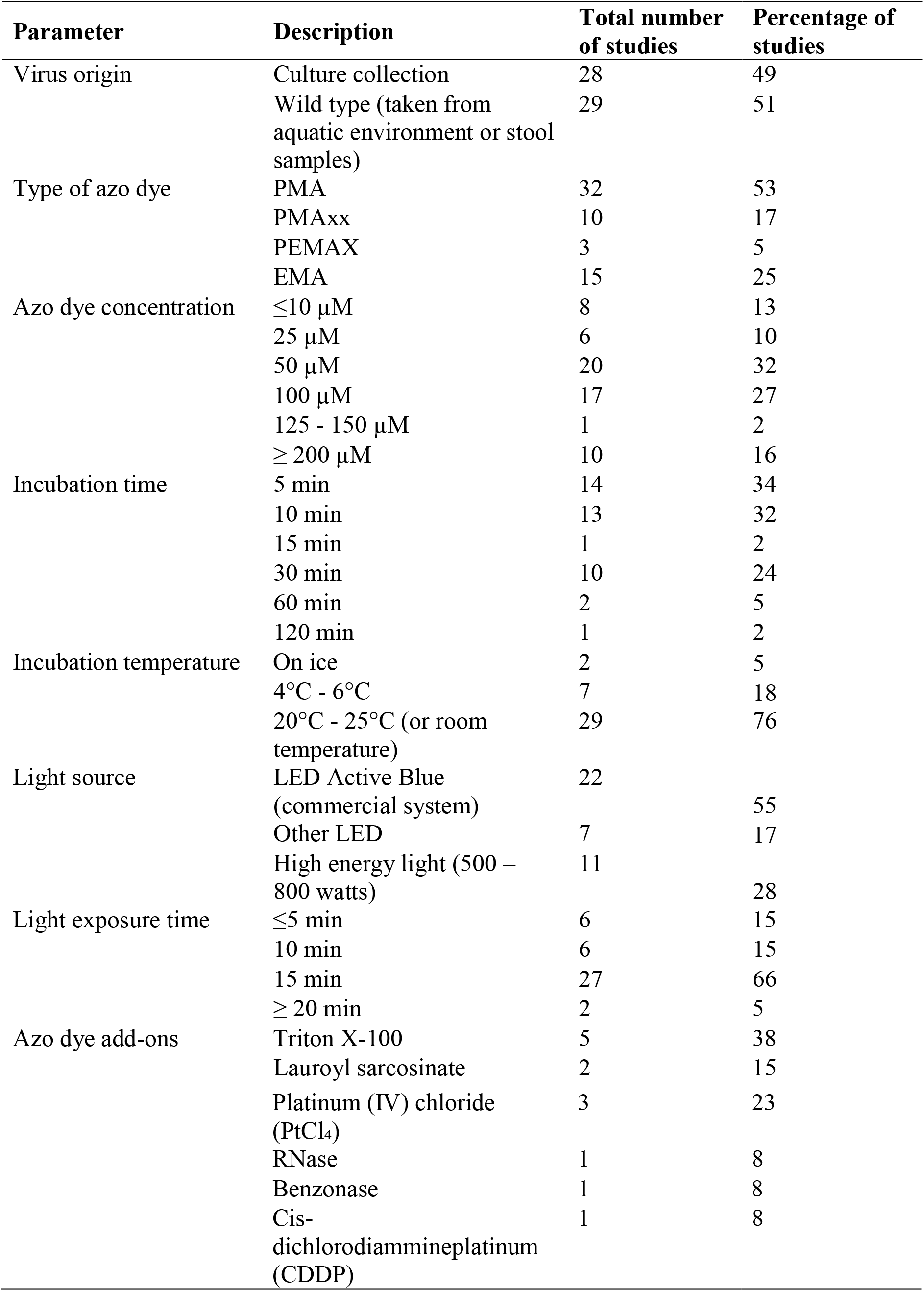
Study design parameters

## 5. Azo dye type and concentration range

Both EMA and PMA can permeate the bacterial cell membrane and virus capsid to intercalate with nucleic acids when activated with light emitted by high-energy lamps or blue light emitting diodes (LEDs). These azo dyes became available for research purposes in the early 2000s, but a number of studies reported that the more charged EMA tended to enter bacterial cells with intact membranes, thus potentially resulting in false-negative qPCR results (Fittipaldi et al., 2012; Gedalanga and Olson, 2009). Seventy percent of the articles we researched used PMA or the derived PMAxx, and a third of the studies compared them to EMA and/or the derivate PEMAX (Table 1). In general, PMA and PMAxx were found to be more suitable for capsid integrity qPCR, most likely due to their higher ability to enter thermally and chemically inactivated cells (Jeong et al., 2017; Kim et al., 2017; Moreno et al., 2015), when compared to infectious virus titers determined by cell culture (Leifels et al., 2019).

Azo dye concentrations are quite evenly distributed, ranging from less than 10 µM to greater than 200 µM. More than half of the articles under investigation reported either 50 µM or 100 µM (Table 1), which is in accordance with the recommendations by the manufacturer for bacterial cultures (Biotium Inc., 2019). However, some works indicated optimal reduction rates with dye concentrations between 4 µM and 10 µM (Karim et al., 2015; Lee et al., 2016; Leifels et al., 2016; Leifels et al., 2019; Prevost et al., 2016). The effect of high azo dye concentration appeared to vary with virus type; for example, adverse effects have been reported with > 125 µM PMA and phage MS2 (Kim and Ko, 2012), while 250 µM PMA worked well for MNV and NoV GII.4 (Jeong et al., 2017; Lee et al., 2015).

## 6. Incubation conditions

Approximately two thirds of the studies included in this review applied an incubation time with the azo dye of 5 or 10 min (Table 1). A quarter incubated for 30 min and less than one in ten of the studies reported periods lasting longer than one hour. A similarly clear trend is observed for incubation temperature; most studies incubated at temperatures between 20°C and 25°C or indicated that room temperature was used (Table 1). The remainder of the articles described the samples being stored on ice or at 4 to 6°C; none of them used higher temperatures as sometimes discussed for bacterial assays (Codony et al., 2019). Studies that applied a long incubation time tended to use low temperatures (Coudray-Mounier et al., 2013; Sangsanont et al, 2014; Prevost et al., 2016; Leifels et al., 2016; Canh et al., 2018; Oristo et al., 2017; Fraisse et al., 2018; Canh et al., 2019; Leifels et al., 2019).

## 7. Dye activation conditions

Early publications utilizing azo dyes for the removal of non-infectious virus particles exclusively applied 500- to 800-Watt halogen light sources used in stage lighting (Bellehumeur et al., 2015; Canh et al., 2018; Escudero-Abarca et al., 2014; Graiver et al., 2010; Leifels et al., 2015; Parshionikar et al., 2010; Sangsanont et al., 2014). Besides their operational hazards such as light bulbs exploding due to long running times (a maximum of 5 min is recommended by the manufacturer), both the heat and light emission in the ultraviolet and infrared spectra could potentially harm the sample, thus subverting the purpose of the pretreatment altogether. Wider availability of consumer-grade LED light technology in general and the introduction of dedicated azo dye activation light sources by companies like GenIUL, Spain, allowed for a much more precise and reproducible activation of PMA, EMA, PEMAX and PMAxx. Not surprisingly, 72% of the records included in this review utilized either the commercial LED Active Blue light system (GenIUL, Spain) or LEDs emitting blue (around 460 nm) light (Lee et al. (2015); Fongaro et al. (2016)) as depicted in Table 1. A similar trend towards uniformity in the protocol design is apparent in the length of time a sample is exposed to the light source. While the operational requirements of the 500- to 800-W halogen lamps severely limited the exposure time to very short periods (2-5 minutes according to manufacturer’s recommendations to avoid excessive heat development that could lead to the light bulb exploding), the standard configuration of the commercial light systems allows for 15 min of intense blue light. Two thirds of the studies therefore employed 15 min as the light exposure time.

## 8. Additional reagents for azo dye pretreatment

Addition of non-ionic surfactants like Triton X-100 (Coudray-Meunier et al., 2013; Moreno et al., 2015) and sodium lauroyl sarcosinate (Lee et al., 2019; Lee et al., 2018) have been described as beneficial for the determination of virus infectivity and were most frequently used in the studies evaluated (Table 1). Inclusion of non-ionic surfactant enable the azo dye molecules to enter partially or completely ruptured capsids, thus improving their binding properties in the virus genome. Palladium and platinum compounds such as Platinum(IV) chloride (PtCl_4_) and Cis-dichlorodiammineplatinum (CPPD) are long known to chelate in mammalian cells by nucleic acid ligands (Rosenberg et al., 1965), and have recently been adopted for the qPCR based discrimination between live and dead bacteria such as *E. coli* and *Cronobacter skazakii* (Soejima et al. (2016). Attempts to use them to determine virus infectivity have been successful but their associated health risk limits potential applications in routine environmental microbiology (Fraisse et al., 2018; Randazzo et al., 2018b).

## 9. Virus inactivation

Various methods to inactivate viruses were used in the studies included in this review. The intention was either to evaluate disinfection efficiency as it is currently used in food safety and water treatment (Jeong et al., 2017; Langlet et al., 2018; Leifels et al., 2016; Randazzo et al., 2018a) or to act as controls to evaluate the efficacy of the capsid integrity protocol (Canh et al., 2018; Canh et al., 2019; Farkas et al., 2020; Leifels et al., 2015). Addition of chlorine, exposure to heat, and proteolysis are known to damage the viral capsid, thus rendering the genome accessible to azo dyes. Temperatures from moderate to high (45°C to 95°C) for ten to thirty minutes could reproducibly demonstrate the ability of all dyes to remove virus signals (to varying degrees) from molecular quantification (Fraisse et al., 2018; Jeong et al., 2017; Leifels et al., 2015; Leifels et al., 2019; Oristo et al., 2018). Similar effects could be shown for the addition of hypochlorite of up to two milligram per milliliter (Fuster et al., 2016; McLellan et al., 2016; Prevost et al., 2016). Light in the ultraviolet spectrum, on the other hand, affects the hydrogen bonds between nucleic acids, resulting in the inability to reproduce inside the host cell. Capsid integrity qPCR failed to capture the loss of virus infectivity in most UV studies (Karim et al., 2015; Kim et al., 2017; Leifels et al., 2016; Leifels et al., 2019); it could detect capsid damage caused by medium-pressure UV lamps, especially at 230-245 nm wavelength (Sangsanont et al., 2014), but not nucleic acid damage caused by other wavelengths in the UV range (Beck et al., 2018; Beck et al., 2014; Sirikanchana et al., 2008a, b). The varying efficiencies of PMA, PMAxx, PEMAX and EMA in preventing the amplification of DNA/RNA of viruses that have lost their ability to infect their host cells due to exposure to heat and reactive substances like chlorine resemble what has been described for bacteria, starting with the first publication on the subject (Nogva et al., 2003).

## 10. Recommendations and potential applications

The utilization of azo dye pretreatment can be recommended for applications where culture assays take too long to inform necessary remedial actions, and under low resource conditions either in developing countries or in laboratories with only basic analytical capabilities. The recent outbreak of SARS-CoV-2 associated COVID-19 represents an example where knowing the ratio of infectious to non-infectious virions would help in determining whether symptomatic or asymptomatic carriers need to be isolated (Beeching et al., 2020; Kaul, 2020). A step-by-step protocol introducing azo-dye pretreatment to determine capsid integrity (and thus virus infectivity) into an established qPCR workflow can be developed (Figure 4) according to the guidelines suggested in this review. The key steps are virus- and matrix-specific optimization of the incubation conditions, dye-concentration and photo activation.

**Figure 4:**
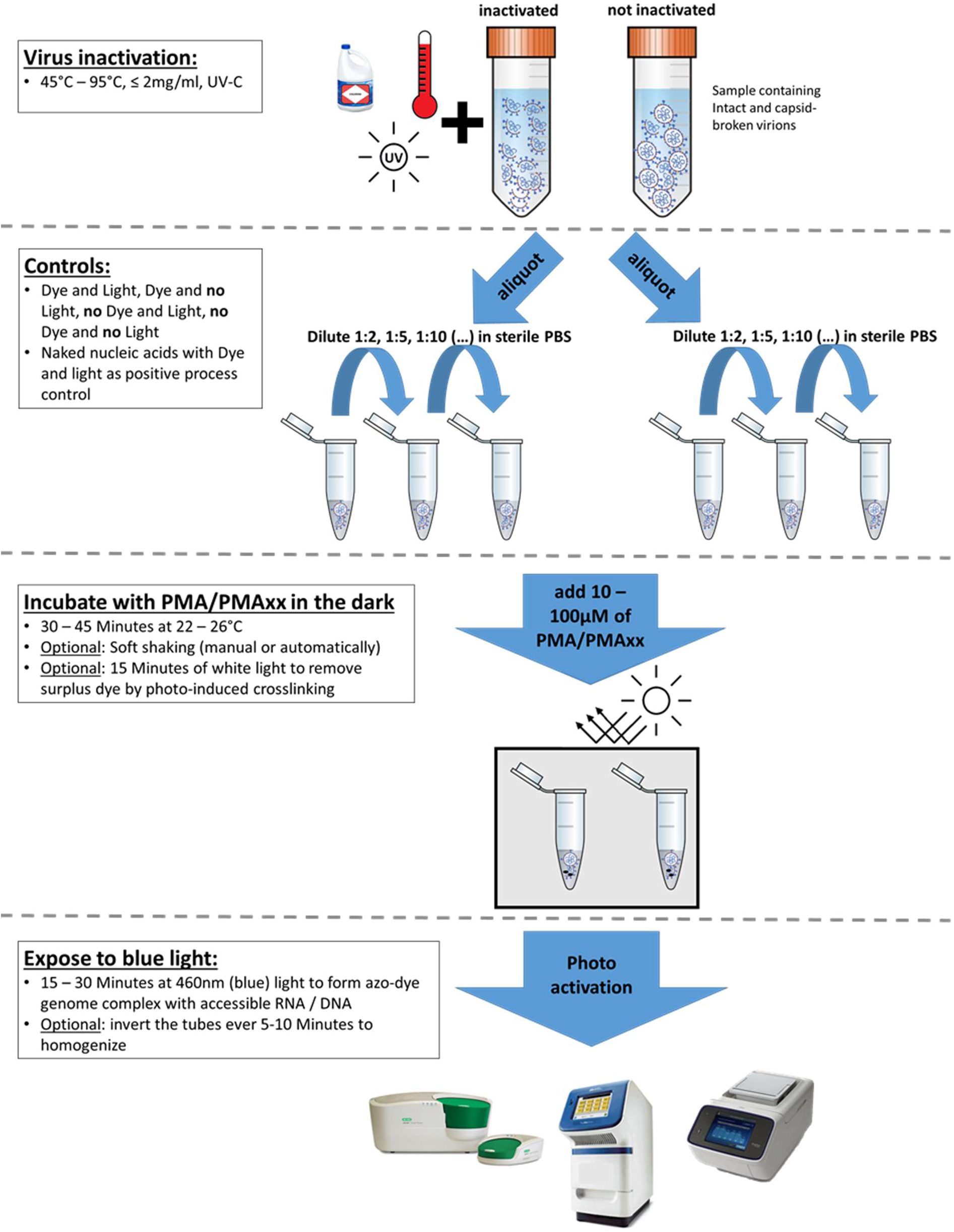
Design of a capsid-integrity qPCR assay. Depending on the sample origin (especially the complexity of the matrix), the virus genus and strain as well as the molecular detection method used, several factors like dilution of the sample, concentration of the azo-dye, incubation conditions and photo activation can be modified and need to be optimized during assay development.

Starting with these initial experimental conditions, optimized protocols need to be generated based on the virus under investigation and the sample origin.

## 11 Conclusions

An evaluation of methods on the application of azo dye pretreatment to determine virus infectivity by qPCR revealed a great diversity of viruses that have been tested under a range of treatment conditions. The systematic literature comparison led to the following conclusions:

- PMA and the derivate PMAxx show a higher efficiency in removing false negative signals from qPCR for both DNA and RNA viruses than EMA and PEMAX.
- Incubation duration and temperature, reagent concentration as well as light source and exposure time need to be optimized and validated for the virus under investigation.
- “One size fits all” pretreatment approaches are possible but might lead to reduced signal reduction rates of individual viruses.
- Capsid integrity qPCR can be a valuable tool to adapt existent workflows and qPCR protocols to reflect the ability of viruses to infect humans, thus improving risk assessment and consumer safety derived from these measurements.
- Capsid integrity is a strong indicator of virus infectivity, which allows for the establishment of robust assays to assess the infectivity of novel viruses in the event of outbreaks like the 2014 Ebola virus epidemic and the current COVID-19 pandemic.

## Data Availability

no data as it is a systematic review

## Author Contributions

M.L. and K. S. conceived and planned the literature review. M. L., E. S. and D. C. S. performed the title and abstract as well as full-text screening. M. L. took lead in writing the manuscript, C. D. and M. L. conceptualized and generated the figures. S. W., S. M. and K. S. provided valuable feedback and helped shape the discussion, analysis and narrative. All authors contributed to editing and proofreading of the manuscript.

## Funding

This research was supported by the Singapore National Research Foundation and Ministry of Education under the Research Centre of Excellence Programme.

## Conflict of interest

The authors declare no conflict of interest.

## Notes

### Competing Interest Statement

The authors have declared no competing interest.

